# Vaccination willingness for COVID-19 among health care workers in Switzerland

**DOI:** 10.1101/2021.07.04.21255203

**Authors:** Kathrin Zürcher, Catrina Mugglin, Matthias Egger, Sandro Müller, Michael Fluri, Laurence Bolick, Rein Jan Piso, Matthias Hoffmann, Lukas Fenner

**Affiliations:** Institute of Social and Preventive Medicine, University of Bern, Switzerland; Kantonsärztlicher Dienst, Gesundheitsamt, Kanton Solothurn, Switzerland; Department of Infectious Diseases, Inselspital, Bern University Hospital, University of Bern, Switzerland; Amt für soziale Sicherheit, Kanton Solothurn, Switzerland; Hausarztpraxis Weissenstein, Langendorf, Switzerland; Department of Internal Medicine, Infectious Diseases and Hospital Epidemiology, Cantonal Hospital Olten, Switzerland

**Keywords:** COVID-19, Vaccine, vaccine hesitancy, health care workers

## Abstract

**Aims of the study:** Vaccination is regarded as the most promising response to the COVID-19 pandemic. We assessed opinions towards COVID-19 vaccination, willingness to be vaccinated, and reasons for vaccination hesitancy among health care workers (HCWs).

**Methods:** We conducted a cross-sectional, web-based survey among 3,793 HCWs in December 2020 in the Canton of Solothurn, Switzerland, before the start of the national COVID-19 vaccination campaign.

**Results:** Median age was 43 years (interquartile range [IQR] 31-53), 2,841 were female (74.9%). 1,511 HCWs (39.8%) reported willingness to accept vaccination, while 1,114 (29.4%) were unsure, and 1,168 (30.8%) would decline vaccination. Among medical doctors, 76.1% were willing, while only 27.8% of nurses expressed willingness. Among 1,168 HCWs who would decline vaccination, 1,073 (91.9%) expressed concerns about vaccine safety and side effects. The willingness of HCWs to be vaccinated was associated with older age (adjusted odds ratio [aOR] 1.97, 95%Cl 1.71-2.27) and having been vaccinated for influenza this year (aOR 2.70, 95%Cl 2.20-3.31). HCWs who reported a lack of confidence in government were less likely to be willing to be vaccinated (aOR 0.58, 95%Cl 0.40-0.84), and women were less willing to be vaccinated than men (OR 0.33 (0.28-0.38).

**Conclusion:** Less than half of HCWs reported willingness to be vaccinated before the campaign start, but proportions varied greatly depending on the profession and workplace. Strategies with clear and objective messages that particularly address the concerns of HCWs are needed if their willingness to be vaccinated is to be increased.

## INTRODUCTION

The worldwide spread of severe acute respiratory syndrome coronavirus 2 (SARS-CoV-2) infections and coronavirus disease 2019 (COVID-19) is a major public health threat (1). Globally, SARS-CoV-2 infection has been confirmed in over 131,837,512 people and as of 7 April, 2021 2,862,664 have died from COVID-19 (2). In Switzerland, 612,575 COVID-19 cases have been confirmed and more than 9,772people have died from COVID-19 (3).

The pandemic has stretched the health care system in Switzerland to its limits and burdened the economy with temporary closures of restaurants and stores and large public and private venues (4). In the absence of effective treatments and a safe and effective vaccine, nonpharmaceutical interventions (NPIs) were implemented to mitigate the pandemic. Measures taken include using personal protective equipment (PPE) such as face masks in public spaces, keeping distance between individuals, and rigorous hand hygiene. Many pharmaceutical companies and research laboratories have been working on vaccines (5-7). Effective vaccination is key to controlling the COVID-19 pandemic, but global vaccine distribution is challenging (8). By the end of 2020, several vaccines had demonstrated efficacy in phase 3 trials (7), and by the beginning of January 2021, two vaccines had been approved by the Swiss Agency for Therapeutic Products (Swissmedic) (9, 10).

The delivery of the COVID-19 vaccines started on 4 January 2021, making it essential to identify and address widespread vaccine uptake barriers. Scepticism about these new vaccines against COVID-19 presents one such challenge to vaccine uptake. Health care workers (HCWs), who face an increased risk of infection with SARS-CoV-2 and can transmit the virus among themselves and to highly vulnerable patients (11-15) are an important target group for vaccination. Previous studies have shown that vaccine uptake for vaccine-preventable diseases such as influenza is low among HCWs (16, 17). Given the central role they play in treating COVID-19 and administering vaccinations, HCWs are uniquely positioned to influence vaccine uptake. Therefore, understanding the willingness of HCWs to be vaccinated against COVID-19, mainly if they are themselves hesitant, will be important in promoting vaccine uptake in the population.

We, therefore, assessed the willingness of healthcare workers in the Canton of Solothurn, Switzerland, to be vaccinated against influenza and COVID-19, and inquired about reasons for vaccine hesitancy among them.

## METHODS

### Study design

We conducted a cross-sectional web-based survey among HCWs in the Canton of Solothurn, Switzerland. We included adults aged 16 or older who work in the health care system in hospitals, medical practices, retirement and nursing homes, home care, pharmacies, and long-term care facilities.

### Data collection

We developed and pilot tested a standardised questionnaire based on the study of Larson et al. (18) and the Vaccine Confidence Project at the London School of Hygiene and Tropical Medicine (www.vaccineconfidence.org). The electronic questionnaire collected information of three types: (i) demographic details such as sex, age, and profession; (ii) intention to be vaccinated against COVID-19 and reasons for being vaccinated/not being vaccinated or for being unsure, confidence in government, recommendation of the employer, and additional information needed to take a vaccination decision; (iii) history of influenza vaccination for influenza season 2020/21. We collected the data in mid-December 2020 before the first approval of a COVID-19 vaccine and associated campaigning using a web-based tool (www.findmind.ch).

All employees of the cantonal hospital in the Canton of Solothurn were invited to participate. The survey was also sent to the cantonal professional associations of physicians, nursing homes, long-term care facilities, and residential care, which invited their members to participate.

### Definitions

The Swiss Federal Office of Public Health (FPOH) defined the following COVID-19 risk groups: persons over the age of 50; those with comorbidities including hypertension, chronic respiratory diseases, diabetes, being immunocompromised, cardiovascular disease, cancer, or obesity (BMI >30); persons who live in a nursing home or long-term care facility; and those who are pregnant.

The survey participants fell into three groups: persons willing to be vaccinated, those who were not willing to be vaccinated, and those who were hesitant or unsure about being vaccinated. Among those willing to be vaccinated against COVID-19, the reasons for deciding to do so could be characterised as self-protection, individual vaccination as a contribution to pandemic control, and membership in a risk group for severe COVID-19 disease. Reasons for unwillingness to be vaccinated included opposition to vaccines in general, the perception that COVID-19 is harmless or that PPE is sufficient, concerns about the effectiveness of the vaccine, its safety and side effects, bad experiences with previous vaccinations, fear of needles, and other reasons. People who were unsure about being vaccinated against COVID-19 were hesitant due to perceived inconsistent information, doubt about vaccine effectiveness, doubt about vaccine safety and fear of side effects, doubt about individual vaccination for pandemic control, and uncertainty among colleagues, and lack of information provided by the employer. Supplementary Table S1 provides further ore details.

### Statistical analyses

We used descriptive statistics to characterise the study population by profession and intention to vaccinate against COVID-19. Differences between groups were assessed using chi square, t-test, or Wilcoxon rank sum tests as appropriate. We calculated the proportions of intention to be vaccinated against COVID-19 with the corresponding 95% confidence intervals (95% CI).

We examined factors associated with the HCW’s intention to be vaccinated against COVID-19 in univariate and multivariate logistic regression. Logistic models were adjusted for age group and profession, confidence in government reports, the employer’s recommendation, and influenza vaccine uptake 2020/21. We grouped uncertainty about vaccination and having no intention of being vaccinated against COVID-19. We also performed a sensitivity analysis that combined those who were willing to be vaccinated and those who were not sure about vaccination. Finally, we compared the vaccine refusers and those who were unsure about COVID-19 vaccination. All analyses were performed in Stata (version 15.1, College Station, TX, USA).

### Ethics statement

Data collection was anonymous. No ethical approval was needed, in line with the Swiss Human Research Act.

## RESULTS

A total of 4,244 HCWs participated in the survey. Response rates were as follows: ∼64% (2,679/∼4,200) among hospital staff, ∼38% (338/∼900) among medical practice staff, ∼11% (440/∼4,000) among nursing home staff, and ∼29% (470/∼1,600) among long term care facility staff.

We excluded participants who did not fully complete the survey (n=421) and those for whom data on sex, age, or profession were missing (n=12). We further excluded participants younger than 16 years (n=18). The analyses thus included 3,793 HCWs (Figure S1).

### Characteristics of the HCWs

The median age of HCW participants was 43 years (interquartile range [IQR] 31-53), 2,841 were female (74.9%). Among all HCWs, 2,445 (64.5%) worked in hospitals, 398 (10.5%) worked in long-term care facilities, 373 (9.8%) in retirement and nursing homes, 311 (8.2%) in medical practices, 232 (6.1%) in residential care, and 34 (0.9%) at pharmacies (Table 1 and Table S2). Asked about COVID-19 vaccination, 1,511 HCWs (39.8%) reported they planned to take the COVID-19 vaccine, 1,114 (29.4%) said they were unsure, and 1,168 (30.8%) said they would not take the vaccination. Willingness to vaccinate was highest among medical doctors, among whom467 of 617 (76.1%) were willing to be vaccinated, and it was lowest among nurses at 470 of 1690 (27.8%) (Figure 1 A). Figure 1 B shows the willingness to be vaccinated by setting.

**Table 1:** Characteristics of HCWs participating in the survey participants, overall and by profession.

|  | All HCWs<br>n=3793 | Nurses<br>n=1690 | Medical<br>doctors<br>n=614 | Medical<br>assistants<br>n=63 | Pharmacist and<br>assistants<br>n=34 | Other medical<br>staff <sup>1</sup><br>n=380 | Private<br>caregivers<br>n=224 | Staff without<br>patient contact<br>n=788 |
| --- | --- | --- | --- | --- | --- | --- | --- | --- |
| **Sex** |  |  |  |  |  |  |  |  |
| Male | 952 (25.1) | 202 (12.0) | 358 (58.3) | 1 (1.6) | 10 (29.4) | 57 (15.0) | 77 (34.4) | 247 (31.4) |
| Female | 2,841 (74.9) | 1,488 (88.0) | 256 (41.7) | 62 (98.4) | 24 (70.6) | 323 (85.0) | 147 (65.6) | 541 (68.7) |
| **Age in years** | 43 (31-53) | 39 (27-52) | 46 (36-55) | 39 (25-52) | 49 (32-59) | 43 (30.5-53) | 41 (30-55) | 48 (36-55) |
| **Workplace** |  |  |  |  |  |  |  |  |
| Hospital | 2,445 (64.5) | 1,122 (66.4) | 372 (60.6) | 0 | 0 | 380 (100) | 0 | 571 (72.5) |
| Medical practice | 311 (8.2) | 6 (0.4) | 240 (39.1) | 58 (92.1) | 0 | 0 | 0 | 7 (0.9) |
| Retirement and nursing home | 373 (9.8) | 287 (17.0) | 0 | 2 (3.2) | 0 | 0 | 0 | 84 (10.7) |
| Residential care | 323 (6.1) | 193 (11.4) | 0 | 2 (3.2) | 0 | 0 | 0 | 37 (4.7) |
| Pharmacy | 34 (0.9) | 0 | 0 | 0 | 34 (100) | 0 | 0 | 0 |
| Long-term care facilities | 398 (10.5) | 82 (4.9) | 2 (0.3) | 1 (1.6) | 0 | 0 | 224 (100) | 89 (11.3) |
| Elderly | - | 0 | 0 | 0 | 0 | 0 | 0 | 0 |
| **Willingness to get COVID-19 vaccine** |  |  |  |  |  |  |  |  |
| Yes | 1,511 (39.8) | 470 (27.8) | 467 (76.1) | 24 (38.1) | 24 (70.6) | 161 (42.4) | 68 (30.4) | 297 (37.7) |
| No | 1,168 (30.8) | 642 (38.0) | 55 (9.0) | 19 (30.2) | 6 (17.6) | 93 (24.5) | 106 (47.3) | 247 (31.3) |
| Unsure | 1,114 (29.4) | 578 (34.2) | 92 (15.0) | 20 (31.8) | 4 (11.8) | 126 (33.2) | 50 (22.3) | 244 (31.0) |
| **Opinion on employer's recommendation** |  |  |  |  |  |  |  |  |
| No opinion | 893 (23.5) | 452 (26.8) | 107 (17.4) | 16 (25.4) | 8 (23.5) | 80 (21.1) | 40 (17.9) | 190 (24.1) |
| Completely and somewhat disagree | 1,066 (28.1) | 598 (35.4) | 75 (12.2) | 17 (27.0) | 3 (8.8) | 86 (22.6) | 95 (42.4) | 192 (24.4) |
| Completely and somewhat agree | 1,834 (48.4) | 640 (37.9) | 432 (79.4) | 30 (47.6) | 23 (67.7) | 214 (56.3) | 89 (39.7) | 406 (51.5) |
| **Opinion on governments reports** |  |  |  |  |  |  |  |  |
| No opinion | 645 (17.0) | 337 (19.9) | 47 (7.7) | 12 (19.1) | 5 (14.7) | 55 (14.5) | 45 (20.1) | 144 (18.3) |
| Completely and somewhat disagree | 868 (22.9) | 489 (28.9) | 48 (7.8) | 18 (28.6) | 6 (17.7) | 57 (15.0) | 71 (31.7) | 179 (22.7) |
| Completely and somewhat agree | 2,280 (60.1) | 864 (51.1) | 519 (84.5) | 33 (52.4) | 23 (67.7) | 268 (70.5) | 108 (48.2) | 465 (59.0) |
| **Had influenza vaccine 20/21** |  |  |  |  |  |  |  |  |
| No | 2,229 (58.8) | 1,101 (65.2) | 113 (18.4) | 34 (54.0) | 11 (32.4) | 217 (57.1) | 193 (86.2) | 560 (71.1) |
| Yes | 1,446 (38.1) | 550 (32.5) | 464 (75.6) | 28 (44.4) | 20 (58.8) | 151 (39.7) | 26 (11.6) | 207 (26.3) |
| Not sure | 118 (3.1) | 39 (2.3) | 37 (6.0) | 1 (1.6) | 3 (8.8) | 12 (3.2) | 5 (2.2) | 21 (2.7) |
| **Reasons for vaccine willingness against COVID-19** | **1,511 (100)** | **470 (100)** | **467 (100)** | **24 (100)** | **24 (100)** | **161 (100)** | **68 (100)** | **297 (100)** |
| Protection | 1,461 (96.7) | 449 (95.5) | 460 (98.5) | 23 (95.8) | 21 (87.5) | 151 (93.8) | 65 (95.6) | 292 (98.3) |
| Vaccine for pandemic control | 1,328 (87.9) | 395 (84.0) | 434 (92.9) | 24 (100) | 21 (87.5) | 143 (88.8) | 55 (80.9) | 256 (86.2) |
| Risk group | 266 (17.6) | 76 (16.2) | 96 (20.6) | 0 | 7 (29.2) | 20 (12.4) | 14 (20.6) | 53 (17.8) |
| **Reasons for vaccine hesitancy against COVID-19** | **1,168 (100)** | **642 (100)** | **55 (100)** | **19 (100)** | **6 (100)** | **93 (100)** | **106 (100)** | **247 (100)** |
| Vaccine safety and side effects | 1,073 (91.9) | 603 (93.9) | 51 (92.7) | 17 (89.5) | 4 (66.7) | 87 (93.5) | 90 (84.9) | 221 (89.5) |
| PPE is sufficient | 328 (28.1) | 193 (30.1) | 13 (23.6) | 4 (21.1) | 1 (16.7) | 26 (28.0) | 32 (30.2) | 59 (23.9) |
| COVID-19 harmless | 229 (19.6) | 118 (18.4) | 20 (36.4) | 3 (15.8) | 0 | 16 (17.2) | 25 (23.6) | 47 (19.0) |
| Bad experiences | 201 (17.2) | 111 (17.3) | 12 (18.2) | 2 (10.5) | 0 | 11 (11.8) | 20 (18.9) | 45 (18.2) |
| Effectiveness of the vaccine | 128 (11.0) | 85 (13.2) | 6 (10.9) | 1 (5.3) | 0 | 8 (8.6) | 12 (11.3) | 16 (6.5) |
| Opponents of vaccines in general | 39 (3.3) | 20 (3.1) | 0 | 1 (5.3) | 0 | 1 (1.1) | 5 (4.7) | 12 (4.9) |
| Fear of needles | 40 (3.4) | 20 (3.1) | 1 (1.8) | 0 | 1 (16.7) | 4 (4.3) | 4 (3.8) | 10 (4.0) |
| Others | 379 (32.4) | 194 (30.2) | 24 (43.6) | 4 (21.1) | 1 (16.7) | 33 (35.5) | 46 (43.4) | 77 (31.2) |
| **Reasons for being unsure of vaccination against COVID-19** | **1,114 (100)** | **578 (100)** | **92 (100)** | **20 (100)** | **4 (100)** | **126 (100)** | **50 (100)** | **244 (100)** |
| Vaccine safety and side effects | 1,058 (95.0) | 538 (93.1) | 88 (95.7) | 18 (90.0) | 4 (100) | 124 (98.4) | 50 (100) | 236 (96.7) |
| Inconsistent information | 329 (29.5) | 192 (33.2) | 29 (31.5) | 1 (5.0) | 1 (25.0) | 40 (31.7) | 17 (34.0) | 51 (20.9) |
| Effectiveness of the vaccine | 316 (28.4) | 177 (30.6) | 16 (17.4) | 5 (25.0) | 1 (25.0) | 33 (26.2) | 15 (30.0) | 69 (28.3) |
| No education by my employer | 210 (18.9) | 126 (21.8) | 14 (15.2) | 5 (25.0) | 0 | 29 (23.0) | 4 (8.0) | 32 (13.1) |
| Uncertainty by my colleagues | 251 (22.5) | 147 (25.4) | 18 (19.6) | 6 (30.0) | 0 | 31 (24.6) | 10 (20.0) | 39 (16.0) |
| Vaccine for pandemic control | 174 (15.6) | 93 (16.1) | 14 (15.2) | 2 (10.0) | 0 | 17 (13.5) | 10 (20.0) | 38 (15.6) |
| **Information needs among unsure HCWs** | **1,114 (100)** | **578 (100)** | **92 (100)** | **20 (100)** | **4 (100)** | **126 (100)** | **50 (100)** | **244 (100)** |
| Information about COVID-19 | 116 (10.4) | 68 (11.8) | 8 (8.7) | 1 (5.0) | 0 | 14 (11.1) | 5 (10.0) | 20 (8.2) |
| Information about the vaccine | 1,055 (94.7) | 550 (95.2) | 81 (88.0) | 19 (95.0) | 4 (100) | 121 (96.0) | 47 (94.0) | 233 (95.5) |
| Reports from vaccinated people | 778 (69.8) | 412 (71.3) | 45 (48.9) | 17 (85.0) | 2 (50.0) | 74 (58.7) | 41 (82.0) | 187 (76.6) |
| Opinion of experts | 404 (36.3) | 220 (38.1) | 32 (34.8) | 9 (45.0) | 0 | 56 (44.4) | 15 (30.0) | 72 (29.5) |

Abbreviations: PPE; personal protective equipment; HCW, health care worker
<sup>1</sup> Other medical staff includes physical therapists, occupational therapists, speech therapists, and midwives

**Figure 1:**
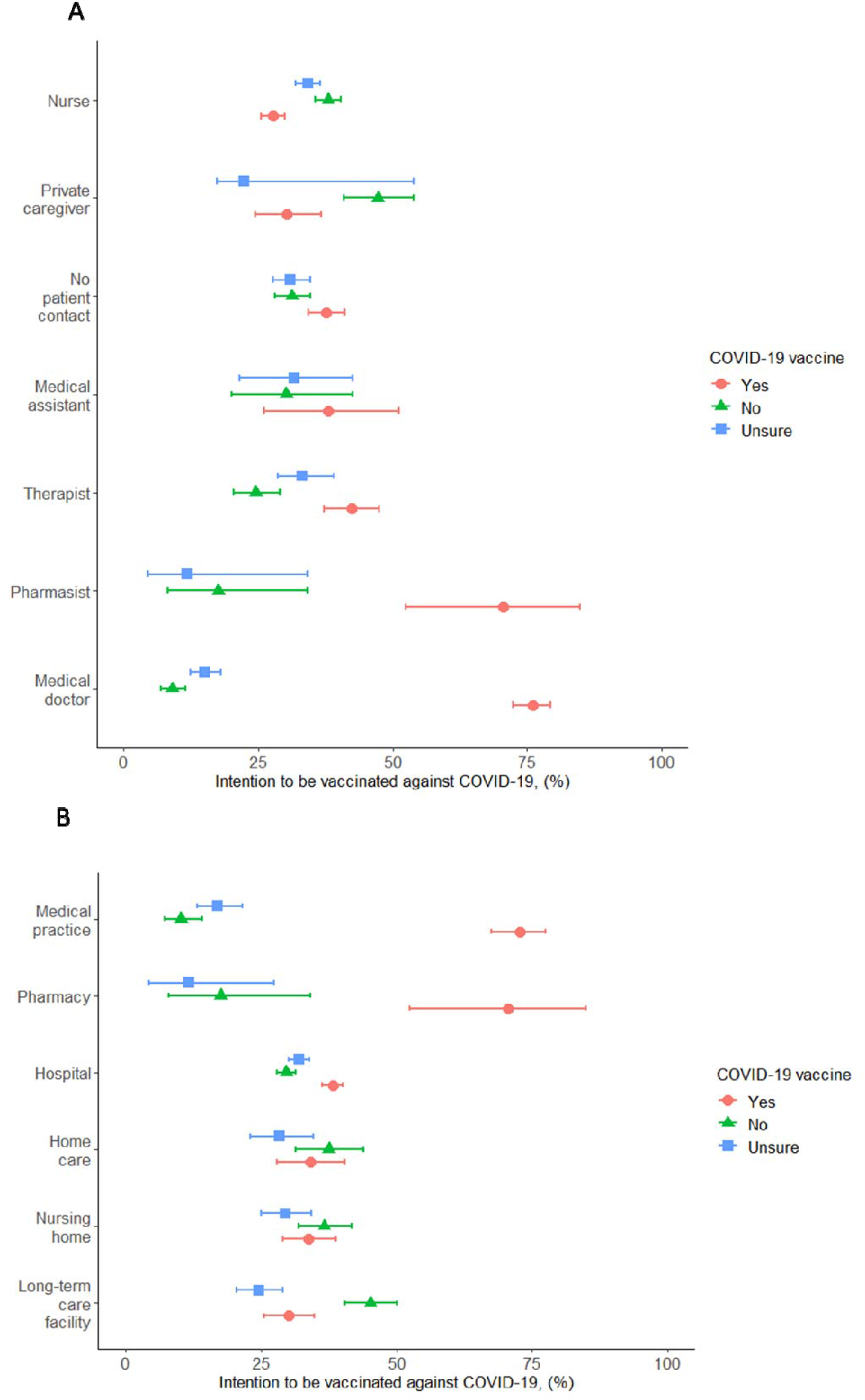
Number of people reporting the willingness to be vaccinated against COVID-19 in the Canton of Solothurn, Switzerland by healthcare profession (A) and by workplace (B). Percent with 95% confidence intervals are presented.

The self-reported uptake for seasonal influenza 2020/2021 vaccination was 38.1% (1,446 of 3,793) while 118 (3.1%) were unsure whether they had been vaccinated, and 2,229 (58.8%) reported they were not vaccinated. The influenza vaccination rate was highest among medical doctors (75.6%) and pharmacists (58.8%). Among the other professions, the vaccination rate ranged from 11.6% to 44.4% (Table 1).

### Determinants of willingness to be vaccinated

The willingness among HCWs to be vaccinated against COVID-19 was lower among females than male participants (OR 0.33, 95%Cl 0.28-0.38) and increased with age. Willingness was highest among the age group ≥ 60 years (OR 4.56, 95%Cl 3.50- 5.94) compared to the age group <30 years. HCWs reporting having been vaccinated against seasonal influenza 2020/21 were more willing to be vaccinated against COVID-19 (OR 6.30, 95%Cl 5.45-7.23) compared to the unvaccinated group. Confidence in government reports and employers’ vaccination recommendation were associated with willingness to be immunised against COVID-19: OR 4.12 (95%Cl 3.37-5.03) and OR 14.19 (95%Cl 11.53-17.47), respectively. In contrast, lack of such confidence was negatively associated with willingness to get vaccinated compared to the group with no opinion (Table 2). The willingness to be vaccinated against COVID-19 was higher among pharmacists (OR 6.22, 95%Cl 2.96-13.13) and medical doctors (OR 8.24, 95%Cl 6.66-12.21) compared to nurses.

**Table 2:** Characteristics of HCWs willingness to get vaccinated against COVID-19, vaccine hesitancy and unsure about vaccination and factors associated with willingness to get vaccinated.

| Variables | Total<br>Number of<br>participants | Willingness to vaccinate against COVID-19 |  |  | Comparing persons with willingness to be vaccinated against Covid-19 to persons with hesitancy/unsure |  |  |  |
| --- | --- | --- | --- | --- | --- | --- | --- | --- |
|  |  | Yes | No | Unsure | unadjusted OR (95% CI) | p-value | adjusted OR (95% CI) * | p-value |
| **Total** | 3,793 | 1,511 | 1,168 | 1,114 |  |  |  |  |
| **Sex** |  |  |  |  |  |  |  |  |
| Male | 952 | 561 (58.9) | 196 (20.6) | 195 (20.5) | 1 | <0.001 |  |  |
| Female | 2841 | 950 (33.4) | 972 (34.2) | 919 (32.4) | 0.33 (0.28-0.38) |  |  |  |
| **Age groups** |  |  |  |  |  | <0.001 |  | <0.001 |
| 29 and younger | 829 | 207 (24.9) | 381 (46.0) | 241 (29.1) | 1 |  | 1 |  |
| 30 to 39 | 807 | 285 (35.3) | 257 (31.9) | 265 (32.8) | 1.64 (1.33-2.03) |  | 0.93 (0.68-1.25) |  |
| 40 to 49 | 790 | 335 (42.4) | 215 (27.2) | 240 (30.4) | 2.21 (1.79-2.73) |  | 1.29 (0.96-1.76) |  |
| 50 to 59 | 1,012 | 470 (46.4) | 254 (25.1) | 288 (28.5) | 2.61 (2.13-3.18) |  | 1.78 (1.34-2.36) |  |
| 60 and older | 355 | 214 (60.3) | 61 (17.2) | 80 (22.5) | 4.56 (3.50-5.94) |  | 2.45 (1.68-3.57) |  |
| **Profession** |  |  |  |  |  | <0.001 |  | <0.001 |
| Medical doctor | 614 | 467 (76.0) | 55 (9.0) | 92 (15.0) | 8.24 (6.66-12.21) |  | 4.88 (3.58-6.65) |  |
| Nurse | 1,690 | 470 (27.8) | 642 (38.0) | 578 (34.2) | 1 |  | 1 |  |
| Medical assistant | 63 | 24 (70.6) | 6 (17.7) | 4 (11.8) | 1.60 (0.95-2.69) |  | 1.45 (0.68-3.10) |  |
| Pharmacist and pharmacist assistant | 24 | 16 (66.7) | 5 (20.8) | 3 (12.5) | 6.22 (2.96-13.13) |  | 4.20 (1.53-11.52) |  |
| Other medical staff <sup>1</sup> | 380 | 161 (42.4) | 93 (24.5) | 126 (33.2) | 1.91 (1.52-2.40) |  | 1.22 (0.89-1.68) |  |
| Not close patient contact | 788 | 297 (37.7) | 247 (31.3) | 244 (31.9) | 1.57 (1.31-1.88) |  | 1.15 (0.89-1.48) |  |
| Private caregiver | 224 | 68 (30.4) | 106 (47.3) | 50 (22.3) | 1.13 (0.83-1.53) |  | 1.51 (0.98-2.33) |  |
| **Confidence in the government reports** |  |  |  |  |  | <0.001 |  | <0.001 |
| No opinion | 645 | 150 (23.3) | 238 (36.9) | 257 (39.8) | 1 |  | 1 |  |
| Completely and somewhat disagree | 868 | 95 (10.9) | 533 (61.4) | 240 (27.7) | 0.41 (0.31-0.54) |  | 0.58 (0.40-0.84) |  |
| Completely and somewhat agree | 2280 | 1,266 (55.5) | 397 (17.4) | 617 (27.1) | 4.12 (3.37-5.03) |  | 1.41 (10.7-1.85) |  |
| **Follow employer's recommendation** |  |  |  |  |  | <0.001 |  | <0.001 |
| No opinion | 893 | 140 (15.7) | 283 (31.7) | 470 (52.6) | 1 |  | 1 |  |
| Completely and somewhat disagree | 1,066 | 41 (3.9) | 801 (75.1) | 224 (21.0) | 0.22 (0.15-0.31) |  | 0.30 (0.20-0.44) |  |
| Completely and somewhat agree | 1,834 | 1,330 (72.5) | 84 (4.6) | 420 (22.9) | 14.19 (11.53-17.47) |  | 11.40 (8.87-14.65) |  |
| **Influenza vaccination 20/21** |  |  |  |  |  | <0.001 |  | <0.001 |
| No | 2,347 | 555 (23.7) | 1019 (43.4) | 773 (32.9) | 1 |  | 1 |  |
| Yes | 1,556 | 956 (66.1) | 149 (10.3) | 341 (23.6) | 6.30 (5.45-7.23) |  | 2.70 (2.20-3.31) |  |

\* Model adjusted for age groups, profession, confidence in the government reports, following employer's recommendation, and influenza vaccination 20/21.
1 Other medical staff includes physical therapists, occupational therapists, speech therapists, and midwives

In the multivariate analysis, willingness to be vaccinated was positively associated with confidence in government reports on the COVID-19 pandemic (aOR 1.59 95%Cl 1.23-2.06) and negatively with lack of confidence in government (aOR 12.85 95%Cl 10.11-16.33). Similarly, confidence in employer recommendations was positively associated, whereas lack of confidence was negatively associated. The analysis confirmed that HCWs who reported vaccination against seasonal influenza 2020/21 were more likely to be willing to be vaccinated against COVID-19 than those who were not vaccinated (aOR 2.70 95%Cl 2.20-3.31) (Table 2).

In the sensitivity analysis grouping the willing and unsure together (rather than the refusers and unsure), results were similar to the primary analysis (Table S3).

### Reasons for vaccine hesitancy

The main reasons for willingness to be vaccinated were personal protection, controlling the pandemic, and belonging to a risk group (Figure 2A, Table 1). These reasons were different by profession (p<0.05). The most frequent reasons for vaccine hesitancy among HCWs included concerns about vaccine safety and side effects (1073/1,168, 91.9%), the perception that personal protective equipment is sufficient (328/1,168, 28.1%), and that COVID-19 is harmless (231/1,168, 19.8% Figure 2B and Table 1). All reasons for vaccine hesitancy were similar in the different HCW professions, except for vaccine safety and side effects, which was mentioned more frequently as a reason for vaccine hesitancy among nurses (p=0.01)

Among 1,114 HCWs who were unsure about a vaccination decision, 1,055 (94.7%) wanted more information on vaccine safety and side effects, 778 (69.6%) awaited reports from already vaccinated people, and 404 (36.3%) wanted an opinion from experts. The main reasons can be found in Figure 2C, Table 1. The reasons for being unsure were similar across the HCW professions, but pharmacists wanted more frequently information on the vaccine than medical doctors (100% vs. 88%, p=0.01).

**Figure 2:**
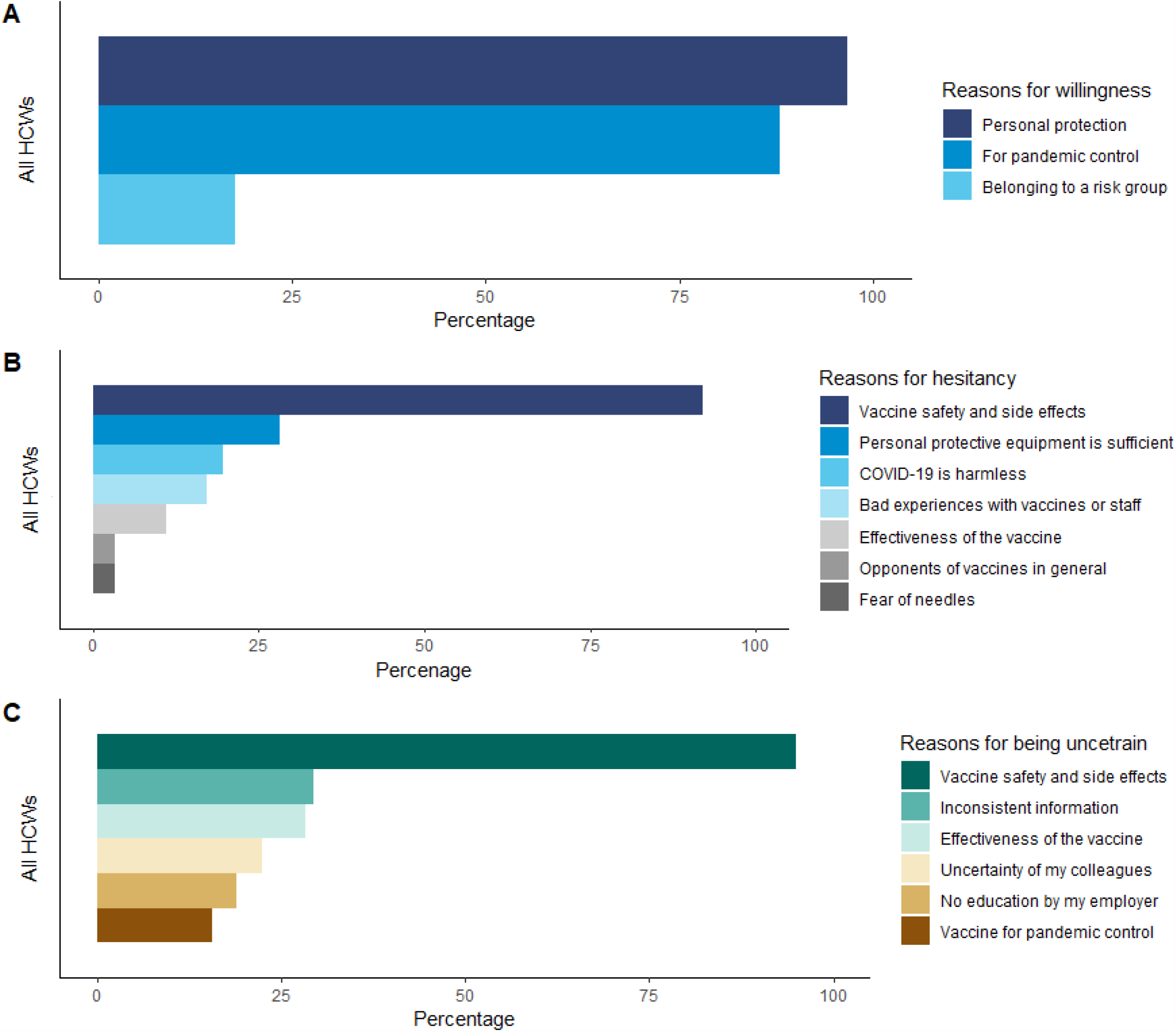
Reasons for vaccine willingness (A), vaccine hesitance (B) and being uncertain (C) to Covid-19 among all HCWs. The exact numbers can be found in Table 1.

## DISCUSSION

Less than half of participating HCWs reported willingness to be vaccinated against COVID-19. The most frequently given reason for vaccine hesitancy was concern about vaccine safety and side effects. Nurses were less likely to be willing to be vaccinated against COVID-19 than were medical doctors and pharmacists.

Vaccines are an effective control measure to reduce the burden of infectious disease. Poliomyelitis was eradicated, and we are close to eliminating measles, mumps, and rubella with vaccines (19). Vaccination will play an important role in the control of the COVID-19 pandemic. Even transient herd immunity in Switzerland will require an estimated 60% of the population to achieve immunity to SARS-CoV-2 either through infection and recovery or vaccination, not considering the potential impacts of SARS-CoV-2 variants of concern. This proportion varies depending on model assumptions (20).

Willingness to get vaccinated is central to achieving herd immunity. Several cross-sectional studies have assessed the willingness to get vaccinated. In seven countries, a European general population survey found that the willingness to be vaccinated against SARS-CoV-2 ranged from 62% to 80% (21). Similar results were found in the United Kingdom and Ireland, with 69% and 65% acceptance, respectively (22). In the USA, 37% of HCWs reported to be willing to get vaccinated against COVID-19 (23). These results are in line with our results (40%) in the Canton of Solothurn in Switzerland. These numbers cause concern because HCWs play an important role in vaccine uptake among the general population. A review on vaccine hesitancy has shown that vaccinated HCWs are more likely to recommend vaccination than unvaccinated HCWs (24). Furthermore, a vaccination recommendation given by an HCW is frequently cited as the reason for vaccination acceptance (25). In contrast, the lack of such a recommendation was the most common reason for not being vaccinated (24, 26).

Some HCWs have expressed reservations about the safety and side effects of COVID-19 vaccines. One contributor to such reservations is the perception of undue haste in the development of COVID-19 vaccines compared to previous vaccines. In the twentieth century, it took a decade or longer to get vaccines to the market, for example, in the case of poliomyelitis (27). Many factors contributed to the comparatively short time it took for COVID-19 vaccines to be granted emergency use authorisations in many countries, with definitive licensing granted on a rolling basis. Researchers have been developing mRNA and viral vector-based vaccines for other diseases for more than a decade (28). As the pandemic spread, public willingness and even demand led to rapid enrolment in phase I through to phase III clinical trials with historically unprecedented speed. Simultaneously, the rapid spread of SARS-CoV-2 allowed endpoint-driven phase III trials to demonstrate vaccine efficacy sooner than was expected at their outset.

We found that confidence in the governmental authorities is associated with willingness to get vaccinated against COVID-19. During a health crisis or a pandemic, trust in the government and risk perception play a key role in vaccine acceptance (29, 30). A French survey among general practitioners showed they were more likely to recommend vaccines to patients when they trusted official sources of information (31). A global survey also reported an association between vaccine acceptance and participants’ trust in government (32). High levels of trust in government was associated with willingness to follow governmental recommendations on preventive behaviors to contain swine flu (33) or compliance with social distancing measures during the Ebola outbreak (34). In contrast, mistrust and misinformation reduced compliance with social and behavioural measures (35). A cross-sectional study during the COVID-19 pandemic among UK residents showed that residents who trusted the government to control the pandemic were more likely to follow government recommendations during the lockdown (36).

Strategies to increase vaccine coverage among HCWs should draw upon guidance from authorities or persons conceived as such by a target audience (in this case HCWs) and specific information about vaccine safety and efficacy. Thus, it is not surprising that many undecided HCWs want to have reports from fellow HCWs who have been vaccinated before they make up their minds. Peers provide important information that influences decision-making, but this source of information is often overlooked. HCWs may be the most effective promotors of vaccination for their fellow workers.

We observed that HCWs who were vaccinated against seasonal influenza are also more likely to willing to be vaccinated against COVID-19. Both diseases are contagious respiratory diseases caused by viruses, and they share some of the same symptoms (fever, cough) and approaches for prevention (hand hygiene, physical distancing, and masks). However, there are important differences. Superspreader events are more common for COVID-19 (37), and mortality rates are higher for COVID-19 than for influenza (38, 39). A recent study showed that over time, the uptake of the seasonal influenza vaccine in Switzerland had dropped overall and among older persons (≥65 years) and people with chronic disease (40). This reduced coverage might be explained by variable and lower seasonal influenza vaccine effectiveness, which can range from 30-60% (41, 42).

The limitations of this study include different response rates among the HCW professions and potential over-representation of hospital-based HCWs. We conducted a web-based cross-sectional online survey in which participation was voluntary but highly supported by the hospital-based personnel. Hospital-based HCWs may be over-represented. HCWs interested in the topic are more likely to respond to the questionnaire, and respondents with biases may select themselves into the sample. This might lead to an overestimation of vaccine hesitancy. Compared to other online surveys, this survey was distributed among specific groups of HCWs to cover this heterogeneous population better. Additionally, attitudes towards vaccination against COVID-19 and willingness to get vaccinated change over time. With more people being vaccinated, acceptance increases, and vaccine hesitancy might decrease.

This study’s strength is the inclusion of diverse health care institutions and HCWs who play quite different roles within them. We included HCWs working with varying populations of risk ranging from nursing home residents, hospitalised patients to people living in long-term care facilities. Furthermore, the survey covered the three largest health care providers in the canton of Solothurn.

## CONCLUSIONS

At this early stage of the COVID-19 vaccine campaign, the overall willingness to get vaccinated among HCWs is low. Balanced and transparent information on vaccine efficacy, safety and side effects should be provided to HCWs and dialogue on vaccine hesitancy initiated.

## Data Availability

All data included in the manuscript.

## AUTHORS CONTRIBUTIONS

Conception and design LF, MH. Study coordination CM, MH, SM. Development of the questionnaire: CM, LF, MH, KZ. Data analysis: KZ, CM, LF, MH wrote the first draft of the paper and revised it based on comments from all authors. All authors reviewed and approved the final version of the manuscript.

## ACKNOWLEDGEMENTS

We would like to thank all HCWs who were willing to participate. Further, we would like to thank all the institutions for distributing the survey to their employees. We thank Christopher Ritter for editorial assistance.

## SOURCES OF SUPPORT

There was no specific funding for this project. KZ was supported by grant U01AI069924 from the U.S. National Institutes of Health’s National Institute of Allergy and Infectious Diseases. ME was supported by special project funding (grant 189498) from the Swiss National Science Foundation.

## COMPETING INTERESTS

All authors declare that they have no conflicts of interest.

## Supplementary Table and Figure

**Table S1:**
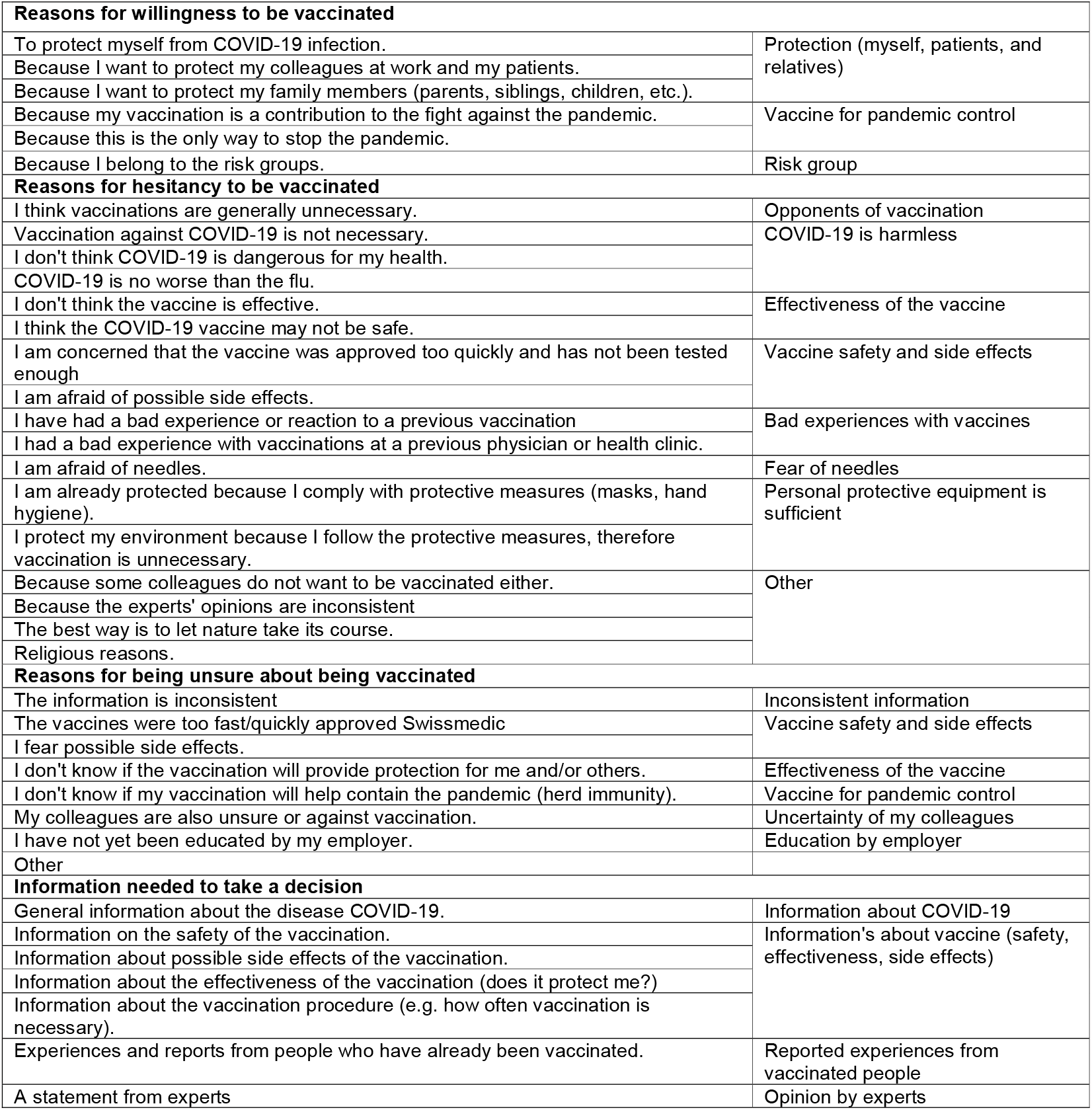
Grouping of the reasons for willingness, hesitancy and unsure to be vaccinated against COVID-19.

**Table S2:** Participant characteristics by workplace.

|  | **Total** | **Hospital** | **Medical practice** | **Nursing home** | **Home care organisation** | **Pharmacy** | **Long-term care facilities** |
| --- | --- | --- | --- | --- | --- | --- | --- |
| **Total** | **3975** | **2445** | **311** | **373** | **232** | **34** | **398** |
| **Sex** |  |  |  |  |  |  |  |
| Male | 952 (25.1) | 576 (23.6) | 164 (52.7) | 59 (15.8) | 15 (6.5) | 10 (29.4) | 128 (6.5) |
| Female | 2841 (74.9) | 1,869 (76.4) | 147 (47.3) | 314 (84.2) | 217 (93.5) | 24 (70.6) | 270 (67.8) |
| **Age in years** | 43 (31-53) | 40 (30-52) | 52 (44-60) | 46 (30-55) | 49.5 (38.5-57) | 49 (32-59) | 44 (31-55) |
| **Workplace** |  |  |  |  |  |  |  |
| Medical doctor | 614 (16.2) | 372 (15.2) | 240 (77.2) | 0 | 0 | 0 | 2 (0.5) |
| Nurse | 1,690 (44.6) | 1,122 (45.9) | 6 (1.9) | 287 (76.9) | 193 (83.2) | 0 | 82 (20.6) |
| Medical assistant | 63 (1.7) | 0 | 58 (18.7) | 2 (0.5) | 2 (0.9) | 0 | 1 (0.3) |
| Other medical staff | 380 (10.0) | 380 (15.5) | 0 | 0 | 0 | 0 | 0 |
| Pharmacist | 34 (0.9) | 0 | 0 | 0 | 0 | 24 (70.6) | 0 |
| Not close patient contact | 788 (20.8) | 571 (23.4) | 7 (2.3) | 84 (22.5) | 37 (16.0) | 0 | 89 (22.4) |
| **Willingness to get COVID-19 vaccine** |  |  |  |  |  |  |  |
| Yes | 1511 (39.8) | 936 (38.3) | 226 (72.7) | 126 (33.8) | 79 (34.0) | 24 (70.6) | 120 (30.2) |
| No | 1,168 (30.8) | 726 (29.7) | 32 (10.3) | 137 (36.7) | 87 (37.5) | 6 (17.6) | 180 (45.2) |
| Unsure | 1,114 (29.4) | 783 (32.0) | 53 (17.0) | 110 (29.5) | 66 (28.5) | 4 (11.8) | 98 (24.6) |
| **Follow my employer's recommendation** |  |  |  |  |  |  |  |
| Completely disagree | 443 (11.7) | 237 (9.7) | 15 (4.8) | 64 (17.2) | 44 (19.0) | 3 (8.8) | 80 (20.1) |
| Somewhat disagree | 623 (16.4) | 403 (16.5) | 32 (10.3) | 61 (16.4) | 47 (20.3) | 0 | 80 (20.1) |
| No opinion | 893 (23.5) | 584 (23.9) | 74 (23.8) | 101 (27.1) | 49 (21.1) | 8 (23.5) | 77 (19.4) |
| Somewhat agree | 742 (19.6) | 528 (21.6) | 41 (13.2) | 54 (14.5) | 43 (18.5) | 6 (17.7) | 70 (17.6) |
| Completely agree | 1,092 (28.8) | 693 (28.3) | 149 (47.9) | 93 (24.9) | 49 (21.1) | 17 (50.0) | 91 (22.9) |
| **Confidence in the governments reports** |  |  |  |  |  |  |  |
| Completely disagree | 230 (6.1) | 129 (5.3) | 7 (2.3) | 37 (9.9) | 14 (6.0) | 0 | 43 (10.8) |
| Somewhat disagree | 638 (16.8) | 388 (15.9) | 33 (10.6) | 74 (19.8) | 48 (20.7) | 6 (17.7) | 89 (22.4) |
| No opinion | 645 (17.0) | 409 (16.7) | 31 (10.0) | 76 (20.4) | 47 (20.3) | 5 (14.7) | 77 (19.4) |
| Somewhat agree | 955 (25.2) | 645 (26.4) | 71 (22.8) | 74 (19.8) | 70 (30.2) | 11 (32.4) | 84 (21.1) |
| Completely agree | 1,325 (34.9) | 874 (35.8) | 169 (54.3) | 112 (30.0) | 53 (22.8) | 12 (35.3) | 105 (26.4) |
| **Influenza vaccine 20/21** |  |  |  |  |  |  |  |
| No | 2,229 (58.8) | 1364 (55.8) | 78 (25.1) | 262 (70.2) | 171 (73.7) | 11 (32.4) | 343 (86.2) |
| Yes | 1,446 (38.1) | 995 (40.7) | 222 (71.4) | 102 (27.4) | 58 (25.0) | 20 (58.8) | 49 (12.3) |
| Unsure | 118 (3.1) | 86 (3.5) | 11 (3.5) | 9 (2.4) | 1 (0.6) | 3 (2.4) | 6 (1.5) |

1 Other medical staff includes physical therapists, occupational therapists, speech therapists, and midwives

**Table S3:** Sensitivity Analysis. Determinants for the willingness to be vaccinated against COVID-19 (yes/unsure) among HCWs compared with HCWs hesitancy.

| Variables | Total<br>Number of<br>participants | Willingness to vaccinate<br>against COVID-19 |  | Comparing the willingness to vaccinate against COVID-19<br>(yes/unsure) to persons with hesitancy |  |  |  |
| --- | --- | --- | --- | --- | --- | --- | --- |
|  |  | Yes | No | unadjusted OR (95%<br>CI) | p-value | adjusted OR<br>(95% CI) * | p-value |
| **Total** | 3,793 | 2,625 | 1,168 |  |  |  |  |
| **Sex** |  |  |  |  | <0.001 |  |  |
| Male | 952 | 756 (79.4) | 196 (20.6) | 1 |  |  |  |
| Female | 2,841 | 1,869 (65.8) | 972 (34.2) | 0.50 (0.42-0.59) |  |  |  |
| **Age groups** |  |  |  |  | <0.001 |  | <0.001 |
| 29 and younger | 829 | 448 (54.0) | 381 (46.0) | 1 |  | 1 |  |
| 30 to 39 | 807 | 550 (68.2) | 257 (31.9) | 1.64 (1.33-2.03) |  | 1.48 (1.12-1.97) |  |
| 40 to 49 | 790 | 575 (72.8) | 215 (27.2) | 2.21 (1.79-2.73) |  | 1.70 (1.23-2.29) |  |
| 50 to 59 | 1012 | 758 (74.9) | 254 (25.1) | 2.61 (2.13-3.18) |  | 2.08 (1.58-2.73) |  |
| 60 and older | 355 | 294 (82.8) | 61 (17.2) | 4.56 (3.50-5.94) |  | 2.72 (1.79-4.13) |  |
| **Profession** |  |  |  |  | <0.001 |  | <0.001 |
| Medical doctor | 614 | 559 (91.0) | 55 (9.0) | 6.22 (4.64-8.35) |  | 1.93 (1.31-2.84) |  |
| Nurse | 1,690 | 1,048 (62.0) | 642 (40.0) | 1 |  | 1 |  |
| Medical assistant | 63 | 44 (69.4) | 19 (30.2) | 1.42 (0.82-2.45) |  | 1.20 (0.56-2.57) |  |
| Pharmacist and pharmacist assistant | 34 | 28 (82.4) | 6 (17.6) | 2.86 (1.18-6.94) |  | 0.85 (0.26-2.79) |  |
| Other medical staff <sup>1</sup> | 380 | 287 (75.5) | 93 (24.5) | 1.89 (1.47-2.44) |  | 1.04 (0.73-1.47) |  |
| Not close patient contact | 788 | 541 (68.6) | 246 (31.4) | 1.34 (1.12-1.61) |  | 0.62 (0.41-0.94) |  |
| Private caregiver | 224 | 118 (52.7) | 106 (47.3) | 0.68 (0.52-0.90) |  | 0.80 (0.62-1.04) |  |
| **Confidence in the governments reports** |  |  |  |  | <0.001 |  | <0.001 |
| No opinion | 645 | 407 (63.1) | 238 (36.9) | 1 |  | 1 |  |
| Completely and somewhat disagree | 868 | 335 (38.6) | 533 (61.4) | 0.37 (0.30-0.45) |  | 0.82 (0.62-1.09) |  |
| Completely and somewhat agree | 2,280 | 1883 (82.6) | 397 (17.4) | 2.77 (2.29-3.36) |  | 1.59 (1.23-2.06) |  |
| **Follow employer's recommendation** |  |  |  |  | <0.001 |  | <0.001 |
| No opinion | 893 | 610 (68.3) | 283 (31.7) | 1 |  | 1 |  |
| Completely and somewhat disagree | 1,066 | 265 (24.9) | 801 (75.1) | 0.15 (0.13-0.19) |  | 0.17 (0.13-0.21) |  |
| Completely and somewhat agree | 1,834 | 1,750 (95.4) | 84 (4.6) | 9.67 (7.45-12.54) |  | 6.58 (4.94-8.76) |  |
| **Influenza Vaccination 20/21** |  |  |  |  | <0.001 |  | <0.001 |
| No | 2347 | 1,328 (56.7) | 1019 (43.4) | 1 |  | 1 |  |
| Yes | 1,556 | 1,297 (89.7) | 149 (10.3) | 6.68 (5.53-8.06) |  | 2.48 (1.94-3.17) |  |

\* Model adjusted for age groups, profession, confidence in the government reports, following employer's recommendation, and influenza vaccination 20/21.
1 Other medical staff includes physical therapists, occupational therapists, speech therapists, and midwives

**Figure S1:**
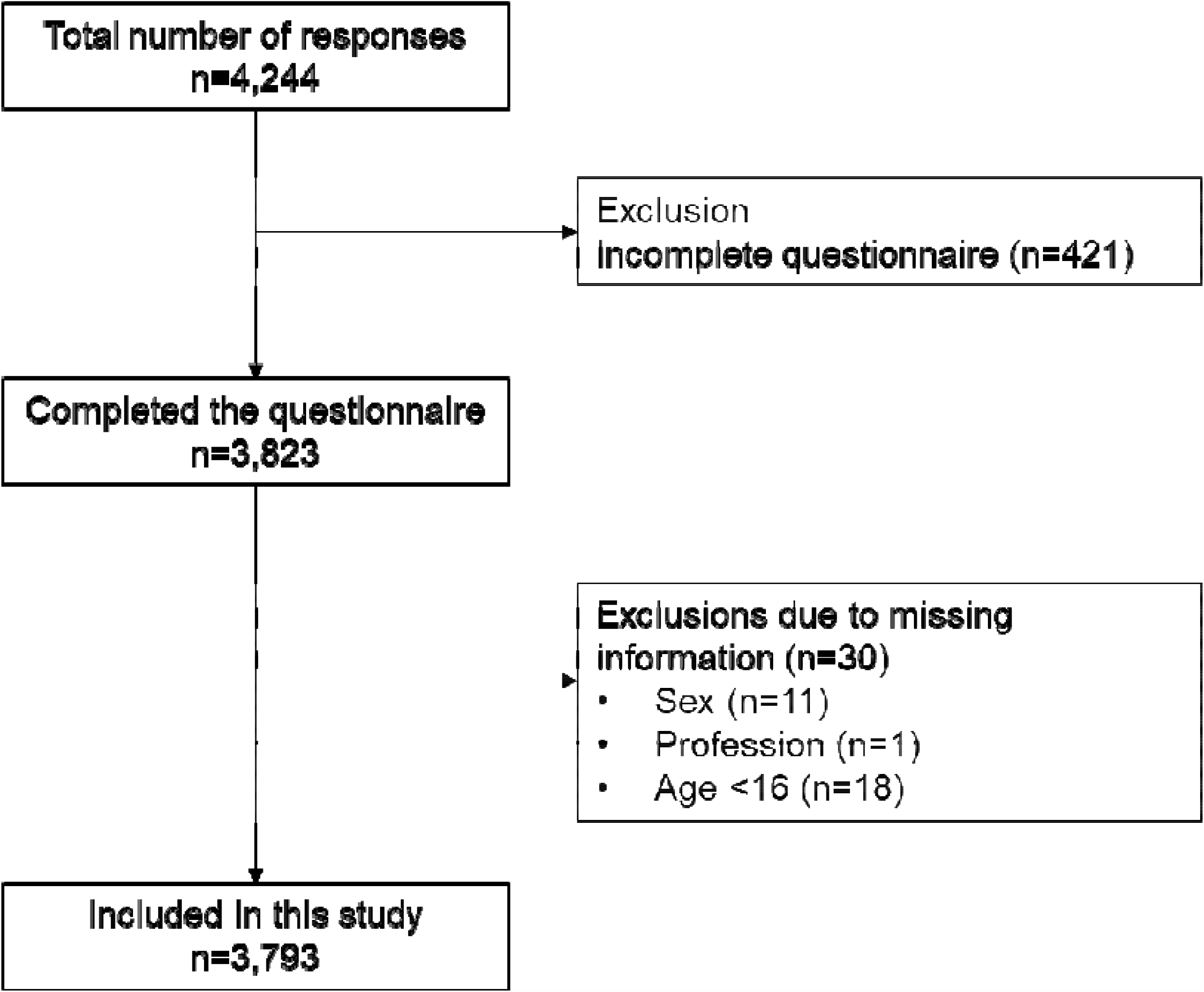
Flow chart.

## Notes

### Competing Interest Statement

The authors have declared no competing interest.

### Author Declarations

Data collection was anonymous. A waiver of ethical approval was obtained from the local Ethics Committee (Cantonal Ethics Committee, EKNZ, Bern, Switzerland) in line with the Swiss Human Research Act.

## REFERENCES

1. Zhu N, Zhang D, Wang W, Li X, Yang B, Song J, et al. A Novel Coronavirus from Patients with Pneumonia in China, 2019. The New England journal of medicine. 2020;382(8):727–33.

2. World Health Organization. Coronavirus Disease (COVID-19) Dashboard Geneva: World Health Organization; 2021 [cited 2021 13 Reburary]. Available from: https://covid19.who.int/.

3. Federal Office of Public Health (FOPH). Coronavirus: Situation Schweiz Bern Federal Office of Public Health; 2021 [Available from: https://www.bag.admin.ch/bag/de/home/krankheiten/ausbrueche-epidemien-pandemien/aktuelle-ausbrueche-epidemien/novel-cov/situation-schweiz-und-international.html.

4. Nicola M, Alsafi Z, Sohrabi C, Kerwan A, Al-Jabir A, Iosifidis C, et al. The socio-economic implications of the coronavirus pandemic (COVID-19): A review. International journal of surgery (London, England). 2020;78:185–93.

5. Kim JH, Marks F, Clemens JD. Looking beyond COVID-19 vaccine phase 3 trials. Nature Medicine. 2021.

6. Parker EPK, Shrotri M, Kampmann B. Keeping track of the SARS-CoV-2 vaccine pipeline. Nature reviews Immunology. 2020;20(11):650.

7. London School of Hygiene & Tropical Medicine. COVID-19 vaccine tracker London: London School of Hygiene & Tropical Medicine; 2020 [Available from: https://vac-lshtm.shinyapps.io/ncov_vaccine_landscape/.

8. So AD, Woo J. Reserving coronavirus disease 2019 vaccines for global access: cross sectional analysis. BMJ. 2020;371:m4750.

9. Swissmedic. Swissmedic grants authorisation for the first COVID-19 vaccine in Switzerland Bern: Swissmedic; 2020 [Available from: https://www.swissmedic.ch/swissmedic/en/home/news/coronavirus-covid-19/covid-19-impfstoff_erstzulassung.html.

10. Swissmedic. Swissmedic grants authorisation for the COVID-19 vaccine from Moderna: Second COVID-19 vaccine authorised in Switzerland Bern: Swissmedic; 2021 [Available from: https://www.swissmedic.ch/swissmedic/en/home/news/coronavirus-covid-19/zulassung-covid-19-impfstoff-moderna.html.

11. Remuzzi A, Remuzzi G. COVID-19 and Italy: what next? The Lancet. 2020.

12. International Council of Nurses. International Council of Nurses High proportion of healthcare workers with COVID-19 in Italy is a stark warning to the world: protecting nurses and their colleagues must be the number one priority. 20 March, 2020. Geneva: International Council of Nurses, ; 2020 [Available from: https://www.icn.ch/news/high-proportion-healthcare-workers-covid-19-italy-stark-warning-world-protecting-nurses-and.

13. Garcia-Basteiro AL, Moncunill G, Tortajada M, Vidal M, Guinovart C, Jiménez A, et al. Seroprevalence of antibodies against SARS-CoV-2 among health care workers in a large Spanish reference hospital. Nature Communications. 2020;11(1):3500.

14. Folgueira MD, Muñoz-Ruipérez C, Alonso-López MÁ, Delgado R. SARS-CoV-2 infection in Health Care Workers in a large public hospital in Madrid, Spain, during March 2020. medRxiv. 2020:2020.04.07.20055723.

15. Thompson HA, Mousa A, Dighe A, Fu H, Arnedo-Pena A, Barrett P, et al. SARS-CoV-2 setting-specific transmission rates: a systematic review and meta-analysis. Clinical infectious diseases : an official publication of the Infectious Diseases Society of America. 2021.

16. Napolitano F, Bianco A, D’Alessandro A, Papadopoli R, Angelillo IF. Healthcare workers’ knowledge, beliefs, and coverage regarding vaccinations in critical care units in Italy. Vaccine. 2019;37(46):6900–6.

17. Harrison N, Brand A, Forstner C, Tobudic S, Burgmann K, Burgmann H. Knowledge, risk perception and attitudes toward vaccination among Austrian health care workers: A cross-sectional study. Human vaccines & immunotherapeutics. 2016;12(9):2459–63.

18. Larson HJ, Schulz WS, Tucker JD, Smith DM. Measuring vaccine confidence: introducing a global vaccine confidence index. PLoS currents. 2015;7.

19. Andre FE, Booy R, Bock HL, Clemens J, Datta SK, John TJ, et al. Vaccination greatly reduces disease, disability, death and inequity worldwide. Bulletin of the World Health Organization. 2008;86(2):140–6.

20. Anderson RM, Vegvari C, Truscott J, Collyer BS. Challenges in creating herd immunity to SARS-CoV-2 infection by mass vaccination. Lancet (London, England). 2020;396(10263):1614–6.

21. Neumann-Böhme S, Varghese NE, Sabat I, Barros PP, Brouwer W, van Exel J, et al. Once we have it, will we use it? A European survey on willingness to be vaccinated against COVID-19. The European journal of health economics : HEPAC : health economics in prevention and care. 2020;21(7):977–82.

22. Murphy J, Vallières F, Bentall RP, Shevlin M, McBride O, Hartman TK, et al. Psychological characteristics associated with COVID-19 vaccine hesitancy and resistance in Ireland and the United Kingdom. Nature Communications. 2021;12(1):29.

23. Shekhar R, Sheikh AB, Upadhyay S, Singh M, Kottewar S, Mir H, et al. COVID-19 Vaccine Acceptance Among Health Care Workers in the United States. medRxiv. 2021:2021.01.03.21249184.

24. Paterson P, Meurice F, Stanberry LR, Glismann S, Rosenthal SL, Larson HJ. Vaccine hesitancy and healthcare providers. Vaccine. 2016;34(52):6700–6.

25. Bianco A, Pileggi C, Iozzo F, Nobile CGA, Pavia M. Vaccination against Human Papilloma Virus infection in male adolescents: Knowledge, attitudes, and acceptability among parents in Italy. Human vaccines & immunotherapeutics. 2014;10(9):2536–42.

26. Smith LE, Amlôt R, Weinman J, Yiend J, Rubin GJ. A systematic review of factors affecting vaccine uptake in young children. Vaccine. 2017;35(45):6059–69.

27. Baicus A. History of polio vaccination. World journal of virology. 2012;1(4):108–14.

28. A gamble pays off in ‘spectacular success’: How the leading coronavirus vaccines made it to the finish line [press release]. Washington, United States The Washington Post 2020.

29. Larson HJ, Clarke RM, Jarrett C, Eckersberger E, Levine Z, Schulz WS, et al. Measuring trust in vaccination: A systematic review. Human vaccines & immunotherapeutics. 2018;14(7):1599–609.

30. van der Weerd W, Timmermans DR, Beaujean DJ, Oudhoff J, van Steenbergen JE. Monitoring the level of government trust, risk perception and intention of the general public to adopt protective measures during the influenza A (H1N1) pandemic in The Netherlands. BMC public health. 2011;11:575.

31. Verger P, Fressard L, Collange F, Gautier A, Jestin C, Launay O, et al. Vaccine Hesitancy Among General Practitioners and Its Determinants During Controversies: A National Cross-sectional Survey in France. EBioMedicine. 2015;2(8):891–7.

32. Lazarus JV, Ratzan SC, Palayew A, Gostin LO, Larson HJ, Rabin K, et al. A global survey of potential acceptance of a COVID-19 vaccine. Nat Med. 2020:1–4.

33. Rubin GJ, Amlôt R, Page L, Wessely S. Public perceptions, anxiety, and behaviour change in relation to the swine flu outbreak: cross sectional telephone survey. BMJ. 2009;339:b2651.

34. Blair RA, Morse BS, Tsai LL. Public health and public trust: Survey evidence from the Ebola Virus Disease epidemic in Liberia. Social Science & Medicine. 2017;172:89–97.

35. Vinck P, Pham PN, Bindu KK, Bedford J, Nilles EJ. Institutional trust and misinformation in the response to the 2018–19 Ebola outbreak in North Kivu, DR Congo: a population-based survey. The Lancet Infectious Diseases. 2019;19(5):529–36.

36. Moxham-Hall V, Strang L. Public opinion and trust in government during a public health crisis London: King’s College London; 2020 [Available from: https://www.kcl.ac.uk/news/public-opinion-and-trust-in-government-during-a-public-health-crisis.

37. Adam DC, Wu P, Wong JY, Lau EHY, Tsang TK, Cauchemez S, et al. Clustering and superspreading potential of SARS-CoV-2 infections in Hong Kong. Nature Medicine. 2020;26(11):1714–9.

38. Piroth L, Cottenet J, Mariet A-S, Bonniaud P, Blot M, Tubert-Bitter P, et al. Comparison of the characteristics, morbidity, and mortality of COVID-19 and seasonal influenza: a nationwide, population-based retrospective cohort study. The Lancet Respiratory Medicine. 2020.

39. Petersen E. COVID-19 is not influenza. The Lancet Respiratory Medicine. 2020.

40. Zürcher K, Zwahlen M, Berlin C, Egger M, Fenner L. Losing ground at the wrong time: trends in self-reported influenza vaccination uptake in Switzerland, Swiss Health Survey 2007–2017. BMJ Open. 2021;11(2):e041354.

41. Sullivan SG, Chilver MB, Carville KS, Deng YM, Grant KA, Higgins G, et al. Low interim influenza vaccine effectiveness, Australia, 1 May to 24 September 2017. Euro surveillance : bulletin Europeen sur les maladies transmissibles = European communicable disease bulletin. 2017;22(43).

42. Sah P, Medlock J, Fitzpatrick MC, Singer BH, Galvani AP. Optimising the impact of low-efficacy influenza vaccines. Proceedings of the National Academy of Sciences of the United States of America. 2018.

